# SARS-CoV-2 RNA in urban wastewater samples to monitor the COVID-19 epidemic in Lombardy, Italy (March – June 2020)

**DOI:** 10.1101/2021.05.05.21256677

**Authors:** Sara Castiglioni, Silvia Schiarea, Laura Pellegrinelli, Valeria Primache, Cristina Galli, Laura Bubba, Federica Mancinelli, Marilisa Marinelli, Danilo Cereda, Emanuela Ammoni, Elena Pariani, Ettore Zuccato, Sandro Binda

## Abstract

Wastewater-based viral surveillance is a promising approach to monitor the circulation of SARS-CoV-2 in the general population. The aim of this study was to develop an analytical method to detect SARS-CoV-2 RNA in urban wastewater, to be implemented in the framework of a surveillance network in the Lombardy region (Northern Italy). This area was the first hotspot of COVID-19 in Europe. Composite 24h samples were collected weekly in eight cities from end-March to mid-June 2020 (first peak of the epidemic). The method developed and optimized, involved virus concentration, using PEG centrifugation, and one-step real-time RT-PCR for analysis. SARS-CoV-2 RNA was identified in 65 (61%) out of 107 samples, and the viral concentrations (up to 2.1 E +05 copies/L) were highest in March-April. By mid-June, wastewater samples tested negative in all the cities. Viral loads were used for inter-city comparison and Brembate, Ranica and Lodi had the highest. The pattern of decrease of SARS-CoV-2 in wastewater was closely comparable to the decline of active COVID-19 cases in the population, reflecting the effect of lock-down. Wastewater surveillance of SARS-CoV-2 can integrate ongoing virological surveillance of COVID-19, providing information from both symptomatic and asymptomatic individuals, and monitoring the effect of health interventions.

## 1. Introduction

The severe acute respiratory syndrome coronavirus-2 (SARS-CoV-2) is responsible for the <u>Co</u>rona <u>Vi</u>rus <u>D</u>isease-19 (COVID-19), a highly infectious respiratory disease that has spread throughout the world at an unprecedented rate from Wuhan, China, since December 2019, affecting over 147 million people and causing more than 3,100,000 deaths so far (ECDC, COVID-19 situation update worldwide). On 11 March 2020, the World Health Organisation (WHO) declared a pandemic and most countries started practising social isolation and restrictions of commercial and industrial activities, which are partially still in place, with enormous socio-economic impact. Healthcare systems in all countries have been – and still are – facing big challenges to tackle the emergency and deal with the need for intensive care for large numbers of patients. In this framework, there is an urgent need for novel tools to monitor the epidemic trends timely and improve the sensitivity and representativeness of public health surveillance systems. Clinical data are currently the main source of information for infectious disease surveillance and tracking, but suffer reporting bias and inability to track asymptomatic carriers (Thompson et al., 2020).

Since the beginning of the pandemic, surveillance of untreated wastewater samples (also called wastewater-based epidemiology - WBE) has been regarded as a promising approach to monitor the circulation of SARS-CoV-2 in the general population (Bivins et al., 2020; Farkas et al., 2020; Lodder and de Roda Husman, 2020). So far, wastewater surveillance has been proposed as a powerful tool for monitoring several factors related to human health, such as the spread of infectious diseases (McCall et al., 2020; Sims and Kasprzyk-Hordern, 2020) and antibiotic resistance (Aarestrup and Woolhouse, 2020; The Global Sewage Surveillance project consortium et al., 2019). Wastewater-based viral surveillance has been used with successful results for early warning of disease outbreaks, informing the efficacy of public health interventions as part of the poliovirus eradication strategy (Lodder and de Roda Husman, 2020; Pellegrinelli et al., 2017), and tracking other enteric viruses such as non-polio enteroviruses, Hepatitis viruses and Rotavirus (Bisseux et al., 2020; Delogu et al., 2018; Fumian et al., 2019; Pellegrinelli et al., 2019). These studies have indicated a good epidemiological link between viral outbreaks and the amounts of viruses in untreated wastewater (sewage), suggesting that wastewater monitoring can complement clinical surveillance of severe diseases giving useful additional information.

It has been demonstrated that SARS-CoV-2 is excreted by faecal route in 30-50% of people who test positive (Parasa et al., 2020; Wang et al., 2020), and it lasts longer in stool than in the respiratory tract (nearly 28 days) (Y. Wu et al., 2020). This means it is likely to be found in sewage samples at the inlet of wastewater treatment plants (WWTPs), composed of the faecal material from an entire community. In fact, SARS-CoV-2 RNA was detected in early March 2020 in wastewater in the Netherlands (Medema et al., 2020), demonstrating for the first time the feasibility of the WBE approach to monitor the circulation of the virus in the population. Wastewater surveillance offers a real-time cost-effective tool for surveying both symptomatic and asymptomatic individuals in an entire community, complementing clinical data where testing is limited to patients with symptoms. Moreover, if applied as routine, it can give early warning of the (re)emergence of SARS-CoV-2 or other viruses in a population, as demonstrated by SARS-CoV-2 detection in wastewater before the first cases were notified to the healthcare systems (La Rosa et al., 2020; Medema et al., 2020).

Wastewater surveillance for SARS-CoV-2 has been applied in studies all over the world: The Netherlands (Medema et al., 2020), Italy (Baldovin et al., 2021; La Rosa et al., 2020; Rimoldi et al., 2020), Spain (Randazzo et al., 2020), Germany (Westhaus et al., 2021), Czech Republic (Mlejnkova et al., 2020), the USA (Gonzalez et al., 2020; Peccia et al., 2020; Sherchan et al., 2020; F. Wu et al., 2020), Australia (Ahmed et al., 2020a), Japan (Haramoto et al., 2020; Hata et al., 2021), United Arab Emirates (Albastaki et al., 2021; Hasan et al., 2021). A global collaborative network was launched in early April 2020 (Bivins et al., 2020) to facilitate the combination of different expertise for undertaking WBE in the fight against COVID-19, through a common platform for data sharing (http://www.covid19wbec.org/) and the Centres for Disease Control and Prevention (CDC) recently published guidance for wastewater-based disease surveillance (CDC, a 2020). A number of countries are applying this approach at a national level as an additional monitoring tool alongside of clinical and epidemiological investigations.

Lombardy was the first hotspot in a country other than China and the outbreak of COVID-19 was one of the deadliest (ECDC, COVID-19 situation update worldwide) including the successive waves of the epidemic. An Italian surveillance strategy called “integrated virological surveillance of COVID-19” was established in response to the epidemic, for tracking infection by collecting respiratory samples mainly from symptomatic individuals (Istituto Superiore di Sanità - ISS). The present investigation aimed to complement this strategy by applying WBE to monitor spatial and temporal profiles of the epidemic by detecting SARS-CoV-2 RNA in urban wastewater collected in Lombardy (Northern Italy) from 31 March (first peak of the pandemic) to mid-June 2020 (end of the first wave). A simple and reliable analytical method was developed and implemented in the framework of a surveillance network in the Lombardy region. This method, based on virus concentration with PEG centrifugation and real-time PCR analysis, was optimized and employed for monitoring the spread of SARS-CoV-2 in the eight cities most affected by COVID-19 in Lombardy. To the best of our knowledge, this is one of the most extensive WBE studies performed for monitoring the presence of SARS-CoV-2 in an area heavily affected by COVID-19.

## 2. Material and methods

### 2.1 Wastewater Sampling

Composite 24h raw wastewater samples were collected at the inlet of 8 Wastewater Treatment Plants (WWTPs) located in 1) Bergamo, 2) Brembate, 3) Ranica, 4) Brescia, 5) Cremona, 6) Crema, 7) Lodi, 8) Milan, in the Lombardy region, North of Italy (Figure 1). Sampling was designed to include the cities most affected by the first wave of COVID-19 in Lombardy, the first area severely hit by COVID-19 in Europe. Figure 1 reports the municipalities investigated with the population served by each WWTP: overall, the combined WWTP catchments resulted in about 2,276,000 inhabitants. As the municipalities affected first and most by the epidemic in Italy, three WWTPs were investigated in the Bergamo province, namely Bergamo, Brembate and Ranica, the last one collecting wastewater from Alzano Lombardo and Nembro. Brescia was another nearby area rapidly involved in the epidemic and the main WWTP of the city was included in the study. Two WWTPs were investigated in Milan covering about 90% of the city population. Finally, three WWTPs in the south of the region were considered, i.e. Lodi, Crema and Cremona, because this was the place where SARS-CoV-2 was first detected and it was the first “red zone” in Italy (the municipality of Codogno).

**Figure 1.**
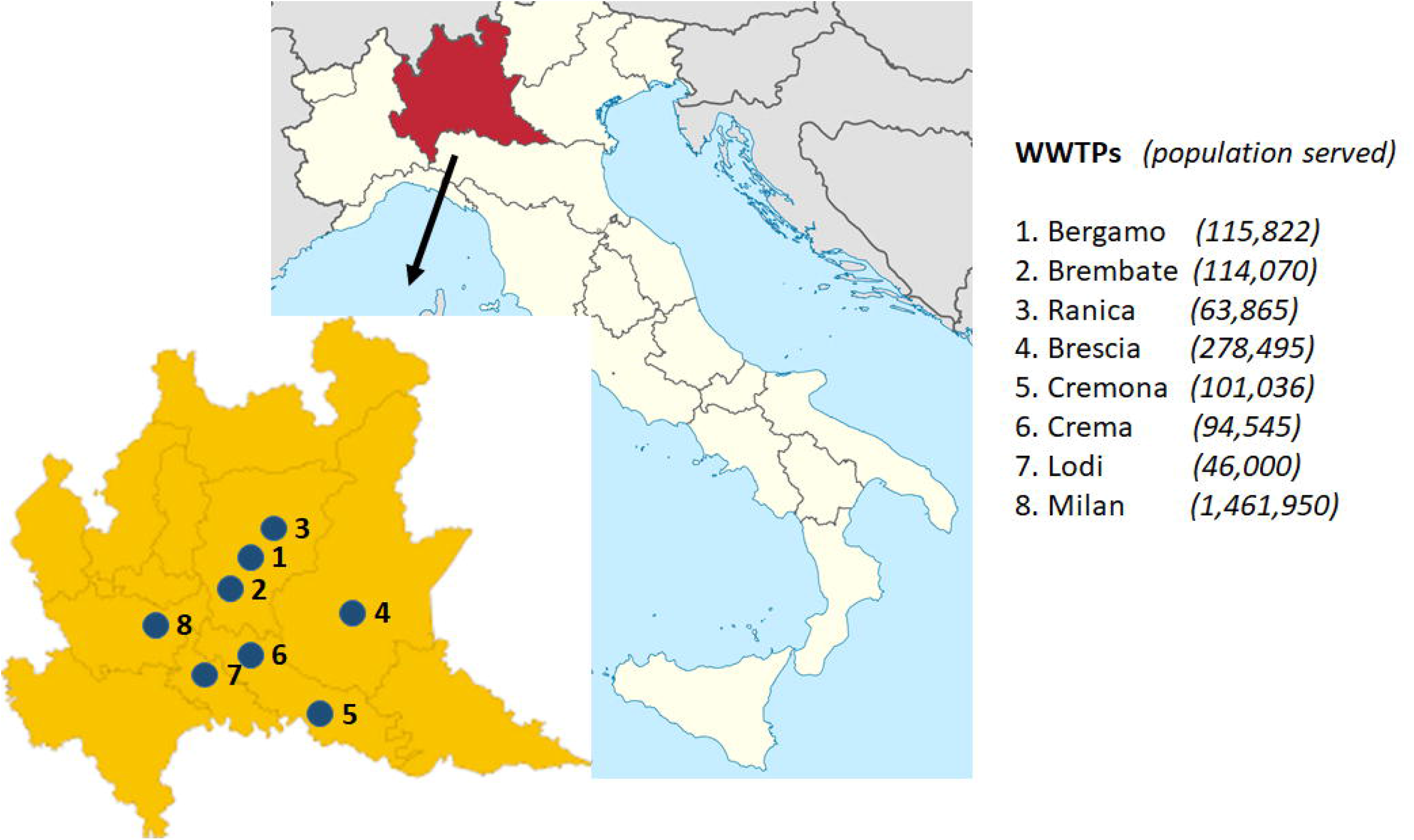
Cities and wastewater treatment plants (WWTPs) investigated in Lombardy, Italy.

Samples were collected once a week from the end of March 2020 to mid-June 2020 in all the sites. Sampling was done in volume or time proportional mode, depending on the automatic sampler available, and following protocols for WBE studies (Castiglioni et al., 2013). Samples were transferred into polypropylene bottles, frozen at −20 °C and transported to the laboratory, where they were stored at the same temperature until analysis (within 4 months).

### 2.2 Concentrations of SARS-CoV-2 from wastewater

Samples were concentrated following the PEG/NaCl centrifugation method proposed by F. Wu et al., 2020, adapted as follows. Raw wastewater (45 mL) was transferred into a 50 mL sterile polypropylene Falcon tube and was left under UV light for 30 minutes to inactivate the infectivity of the virus. Two tubes per sample were processed (total volume 90 mL) and were kept under rotation during UV exposure. Thermic inactivation was also tested at different conditions, among which those suggested in the literature (60°C for 30 minutes)(F. Wu et al., 2020), and gave comparable results to the UV light treatment. Samples were centrifuged for 30 min at 4,500 x g and 4°C, without brake, to remove particulate biomass (Centrifuge Eppendorf 5810 R). After centrifugation, 40 mL of supernatant were transferred into a fresh Falcon tube containing 4 g PEG 8000 and 0.9 g sodium chloride (Carlo Erba, Italy), and left in a shaker for 15 min at room temperature to dissolve the PEG. Samples were then centrifuged for 2h at 12,000 x g and 4 °C, without brake. After centrifugation, the supernatant was discarded and the tubes were returned to the centrifuge at 4°C for a second step at 12,000 x g for 5 min. The pellet in each tube was suspended in 750 µL of tryzol (Life Technologies, Italy); the two aliquots per sample were merged and stored at −20 °C until RNA extraction.

### 2.3 RNA extraction and SARS-CoV-2 molecular detection

Viral RNA was extracted from sewage concentrates using the commercial kit QIAamp MinElute Virus Spin Kit (Qiagen, Germany) according to the manufacturer’s instructions for large samples. Briefly, a wastewater sample concentrate (volume optimized 400 µL) was added to 450 µL of lysis mixture containing Buffer AL, carrier RNA and proteinase K. The lysate was added to 500 µL of ethanol (96-100%) and nucleic acids were bound to the silica membrane of the QIAamp MinElute column by centrifugation. After three washing steps, nucleic acids were eluted in buffer AVE. Extraction was done using the automatic extractor QIAcube (Qiagen, Germany).

SARS-CoV-2 RNA was detected by a one-step real-time RT-PCR assay, amplifying different portions of the nucleocapsid (N) gene, namely N1 and N3, in accordance with the CDC protocol (CDC, b 2020). Specific positive (EURM-019) and negative (DNAse/RNAse-free distilled water) controls (three to six) were tested in all real-time RT-PCR runs.

To semi-quantify the SARS-CoV-2 viral load, a standard curve was plotted using 10-fold serial dilutions of the positive control material (EURM-019) provided by the European Commission. A sample was considered positive for SARS-CoV-2 when N1 or N3 or both viral targets showed a Cycle threshold (Ct) <39. Real-time RT-PCR runs were done with the QuantStudio 5 Real-time RT-PCR system (ThermoFisher Scientific, USA).

To assess the sensitivity of the different SARS-CoV-2 extraction protocols, results from a droplet digital PCR (ddPCR) was appraise. Concentrated wastewater samples were prepared using a droplet generator QX200 AutoDG (Bio-Rad, California, USA) and analysed with a QX200 ddPCR and droplet reader (Bio-Rad, California, USA). The 2019-nCoV CDC ddPCR Triplex Probe Assay was used to quantify the viral genome by targeting two regions of the N gene (N1 and N2). The PCR protocol was the one suggested by Bio-Rad (Bio-Rad SARS-CoV-2 ddPCR Kit) and the sample volume loaded for each PCR reaction was 5.5 µL. Samples were analysed in triplicate and controls (three to six) were run within each batch using RNA-free water.

To minimize potential Real-time RT-PCR contamination, RNA extraction, molecular assays set-up and real-time RT-PCR runs were all done in separate laboratories.

### 2.4 Optimization of viral concentration and RNA extraction

Different protocols for viral RNA concentration were tested to choose the most efficient for concentration from wastewater samples. A wastewater sample positive to SARS-CoV-2 in a preliminary investigation was treated using three different procedures: i) the two-phase (PEG/dextran) method from the 2003 WHO Guidelines for Environmental Surveillance of Poliovirus protocol, modified for enveloped viruses (La Rosa et al., 2020); ii) the PEG/NaCl centrifugation method (centrifuge speed 12000 x g) modified from F. Wu et al., 2020; iii) the PEG/NaCl centrifugation method modified using PEG 6000 and PEG 8000 at a lower centrifugation speed (3890 x g). Details of each protocol are reported in the Supplementary Material (SM).

Viral RNA extraction was optimized by testing different volumes of concentrate: i) 200 µL and ii) 400 µL and different elution volumes: i) 100 µL, ii) two consecutive 50 µL elutions (as suggested by the manufacturer) for a total of 100 µL, and iii) 60 µL. To increase the sensitivity of analysis and have quantitative results, the droplet digital PCR (ddPCR) was used for these optimization tests.

### 2.5 Quality control of the optimized protocol

Once an optimal method of sewage concentration and extraction procedure was set, a quality control experiment was run to check the recovery rate of SARS-CoV-2 RNA and determine the presence of real-time RT-PCR inhibition. A supernatant of cell culture infected by SARS-CoV-2 with a viral load of 2.66E+12, was diluted 1:1000 to obtain a viral load of 2.66E+9 and added to a sample of wastewater, followed by a pasteurization procedure to inactivate the replication of the virus (56° for 1 h). After concentration, the spiked wastewater sample was analyzed in triplicate to quantify SARS-CoV-2 RNA recovery molecularly.

### 2.6 Data analysis

SARS-CoV-2 RNA concentrations, obtained as copies of RNA out of 1 liter of wastewater, were multiplied by the daily flow rate of each WWTP (m^3^/day) to obtain the viral loads entering the WWTPs daily from the population served. Loads were then normalized to the population to compare results from the different communities. The residential population was available for almost all the WWTPs and was used for normalization, in other cases the population equivalents estimated using hydrochemical parameters (i.e. biological oxygen demand - BOD) were used following a best practice protocol for data normalization (Castiglioni et al., 2013).

Viral loads were then compared to the number of “COVID-19 Active cases” at the date of sampling, estimated by summing the new cases diagnosed daily in the previous four weeks. The daily numbers of new cases in the different municipalities were obtained from The Economist (GitHub) and from the Regione Lombardia database (Regione Lombardia, 2020). A four week period was used considering the extended fecal excretion of SARS-CoV-2 up to 28 days after diagnosis, reported in the literature (Y. Wu et al., 2020).

## 3. Results and Discussion

### 3.1 Protocol optimization

The wastewater samples analysed using PEG/dextran procedure (i) were negative, while those analysed with the PEG/NaCl centrifugation methods (ii and iii) gave positive results, with the best performance using PEG 8000 and centrifuge at 12000xg (Table S1). This best protocol was also tested by adding the particulate collected from the first centrifuge to the water concentrate before RNA extraction (see SM for details), and performance improved (Table S1), though further investigation is needed to confirm this result.

The best volume for viral RNA extraction was 400 µL (Table S2), and ddPCR clearly showed a double number of gene copies/mL when using 400 µL instead of 200 µL (Table S2). Considering the elution volume, the best result was obtained with 60 µL (Table S3). The limit of detection (LOD) of the amplification assay with the optimized concentration and extraction methods was 1 copy/µL.

### 3.2 Quality control of the extraction procedure

After sample concentration, the viral load of SARS-CoV-2 in spiked wastewater was 2.5E+9 (Ct 20.1) with a variability of replicates (relative standard deviation - RSD) of 0.4% (Table 1). The repeatability of the optimized extraction procedure for triplicate analysis of a real (not spiked) wastewater sample was good, with a RSD of 1%. The amount of SARS-CoV-2 recovered was fully comparable with the amount spiked in the wastewater samples (viral load 2.66E+9).

**Table 1.**
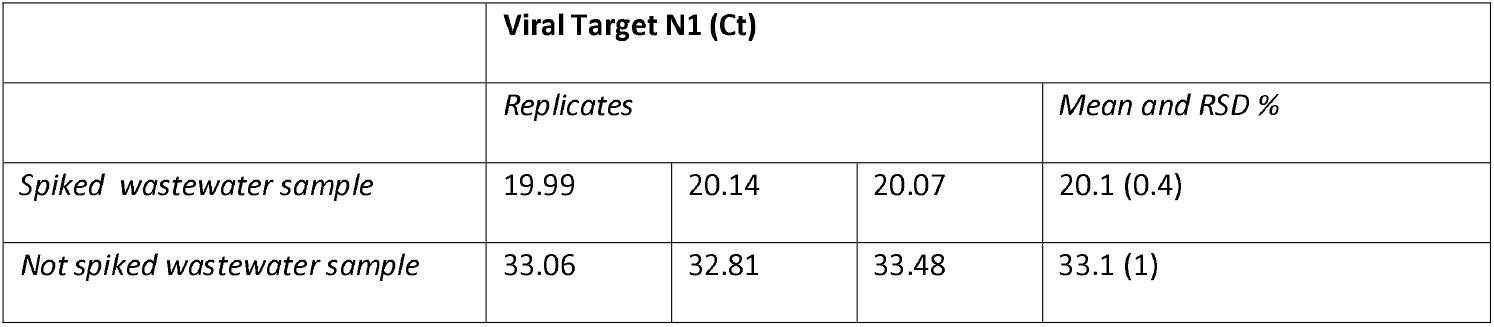
Extraction efficiency of SARS-CoV-2 from spiked wastewater samples and repeatability for triplicate analysis of a real sample.

These results confirmed the efficiently of viral recovery from the optimized procedure and the absence of real-time RT-PCR inhibitors.

### 3.3 Detection of SARS-CoV-2 RNA in wastewater samples

At the end of February 2020, Italy reported the first two clusters of autochthonous SARS-CoV-2 infection, identified in the provinces of Lodi and Bergamo (Codogno and Alzano Lombardo municipalities, respectively) in Lombardy. This region was the most affected with 94,000 COVID-19 cases and over 16,500 related deaths up to June 2020 (Regione Lombardia, Dashboard COVID-19). The national lock-down with schools, offices, restaurants, bars and shops closed excluded those for primary needs such as food and medical supplies, and with people at home, was obligatory from mid-March 2020. The present study was conducted from the end of March to mid-June 2020, in the midst of the first Italian wave of COVID-19, starting two weeks after the national lock-down. Wastewater samples were collected at the inlet of 8 WWTPs in the Lombardy region and were frozen at −20 °C after collection; this procedure did not affect the SARS-CoV-2 concentration since freezing temperatures can be used for storage when immediate SARS-CoV-2 RNA analysis from the wastewater influent is not possible (Hokajärvi et al., 2021).

SARS-CoV-2 RNA was identified in 65 out of 107 (61%) wastewater samples collected in Lombardy (Figure 1). The rate of positive samples was highest in the Bergamo area (78.7%; 26/33), with 83.3% (7/11) of positive samples in Brembate, 75% (9/12) in Ranica, and 63.6% (7/11) in Bergamo. A similar rate of positive samples was found in the close area of Brescia (77.8%; 7/9). Positive samples were slightly lower in the other areas, where respectively, 66.7% (8/12) and 61.5% (16/26) of sewage samples from WWTPs in Lodi and Milan tested positive for SARS-CoV-2, and 58.3% (7/12) in Cremona and Crema. As reported in Tables 2–4, the temporal trends of SARS-CoV-2 circulation across Lombardy were similar; SARS-CoV-2 was highest in March-April 2020, followed by a steep decline, with samples starting to give negative results for SARS-CoV-2 at the end of May, as observed in the Veneto region, close to Lombardy (Baldovin et al., 2021).

**Table 2.** SARS-CoV-2 in wastewater in north-western Lombardy (Bergamo and Brescia provinces). Viral targets were two different portions of the nucleocapsid (N) gene, N1 and N3 (copies/L).

| Bergamo WWTP |  |  | Brembate WWTP |  |  | Ranica WWTP |  |  | Brescia WWTP |  |  |
| --- | --- | --- | --- | --- | --- | --- | --- | --- | --- | --- | --- |
| *Sampling*<br>*dates* | *N1*<br>*copies/L* | *N3*<br>*copies/L* | *Sampling*<br>*dates* | *N1*<br>*copies/L* | *N3*<br>*copies/L* | *Sampling*<br>*dates* | *N1*<br>*copies/L* | *N3*<br>*copies/L* | *Sampling*<br>*dates* | *N1*<br>*copies/L* | *N3*<br>*copies/L* |
| 30/03/2020 | 1,02E+04 | 1,03E+04 | 30/03/2020 | 2,10E+05 | 1,22E+05 | 01/04/2020 | 2,28E+04 | 1,56E+04 | 30/03/2020 | 6,28E+04 | 3,98E+04 |
| 05/04/2020 | 7,29E+03 | 4,95E+03 | 06/04/2020 | 9,73E+04 | 7,93E+04 | 05/04/2020 | 2,58E+04 | 1,69E+04 | 06/04/2020 | 2,22E+04 | 1,36E+04 |
| 13/04/2020 | 4,61E+03 | 4,04E+03 | 14/04/2020 | 1,97E+04 | 2,12E+04 | 08/04/2020 | 6,24E+03 | 1,38E+04 | 13/04/2020 | 7,03E+03 | 8,24E+03 |
| 21/04/2020 | 2,06E+03 | 1,86E+03 | 22/04/2020 | 1,67E+04 | 1,76E+04 | 15/04/2020 | 5,00E+03 | 1,95E+04 | 21/04/2020 | 3,81E+03 | 5,57E+03 |
| 29/04/2020 | 1,42E+03 | nd | 29/04/2020 | 1,50E+03 | 1,99E+03 | 19/04/2020 | 8,91E+03 | 8,61E+03 | 28/04/2020 | 2,53E+03 | 4,51E+02 |
| 04/05/2020 | 1,06E+03 | 2,65E+03 | 06/05/2020 | 3,37E+03 | 3,59E+03 | 26/04/2020 | 3,24E+03 | 4,03E+03 | 12/05/2020 | nd | nd |
| 12/05/2020 | 1,15E+03 | nd | 13/05/2020 | nd | 4,55E+03 | 03/05/2020 | 9,45E+03 | 1,10E+04 | 19/05/2020 | 2,72E+03 | nd |
| 19/05/2020 | nd | nd | 20/05/2020 | 1,51E+03 | 2,31E+03 | 10/05/2020 | nd | nd | 26/05/2020 | nd | nd |
| 27/05/2020 | nd | nd | 27/05/2020 | 3,82E+03 | 5,51E+03 | 17/05/2020 | 3,57E+03 | 4,11E+03 | 16/06/2020 | nd | 2,70E+03 |
| 10/06/2020 | nd | nd | 03/06/2020 | 1,39E+03 | 4,63E+03 | 24/05/2020 | nd | nd |  |  |  |
| 16/06/2020 | nd | nd | 10/06/2020 | nd | nd | 07/06/2020 | nd | nd |  |  |  |
|  |  |  | 17/06/2020 | nd | nd | 14/06/2020 | nd | 3,84E+03 |  |  |  |

Overall, the highest SARS-CoV-2 concentrations were in the samples collected in late March and early April. From the province of Bergamo, the highest SARS-CoV-2 concentration was in Brembate (2.10+05 copies/L), followed by Ranica (2.28E+04 copies/L) and Bergamo (1.02E+04 copies/L) at the end of March (Table 2). The SARS-CoV-2 genome was identified until mid-May in Bergamo and Ranica and until mid-June in Brembate. In the Brescia WWTP, the concentration of SARS-CoV-2 was highest at the end of March (6.28+04 copies/L) and the virus was detected until early May 2020 (Table 2). In the south of Lombardy, Lodi was the most affected province, where the virus was detected until mid-May with the highest concentration of SARS-CoV-2 for N1 assay at the end of March (7.64 E+04 copies/L) (Table 3). In Cremona and Crema, SARS-CoV-2 circulated until early May with the highest concentrations in early April (8.16 E+03 copies/L and 6.96 E+03 copies/L, respectively) (Table 3).

**Table 3.** SARS-CoV-2 in wastewater in the south of Lombardy (Cremona and Lodi provinces). Viral targets were two different portions of the nucleocapsid (N) gene, N1 and N3 (copies/L).

| Cremona WWTP |  |  | Crema WWTP |  |  | Lodi WWTP |  |  |
| --- | --- | --- | --- | --- | --- | --- | --- | --- |
| *Sampling dates* | *N1 copies/L* | *N3 copies/L* | *Sampling dates* | *N1 copies/L* | *N3 copies/L* | *Sampling dates* | *N1 copies/L* | *N3 copies/L* |
| 31/03/2020 | 8,16E+03 | 9,15E+03 | 31/03/2020 | 6,96,E+03 | 6,85,E+03 | 30/03/2020 | 7,64E+04 | 8,29E+04 |
| 05/04/2020 | 9,19E+03 | 5,48E+03 | 07/04/2020 | 1,47,E+04 | 1,27,E+04 | 01/04/2020 | 9,29E+03 | 1,01E+04 |
| 10/04/2020 | 1,02E+04 | 3,20E+03 | 14/04/2020 | 9,91,E+03 | 3,21,E+03 | 05/04/2020 | 8,24E+03 | 1,26E+04 |
| 16/04/2020 | 2,39E+03 | 1,90E+03 | 21/04/2020 | 1,21,E+03 | 5,30,E+03 | 13/04/2020 | 1,62E+04 | 1,93E+04 |
| 23/04/2020 | 2,00E+03 | 3,61E+03 | 28/04/2020 | 2,78,E+03 | 6,25,E+03 | 21/04/2020 | 8,44E+03 | 1,14E+04 |
| 28/04/2020 | 2,02E+03 | 1,52E+03 | 05/05/2020 | 2,54,E+03 | nd | 28/04/2020 | 1,53E+04 | 1,22E+04 |
| 05/05/2020 | nd | nd | 12/05/2020 | nd | nd | 05/05/2020 | nd | nd |
| 12/05/2020 | nd | nd | 20/05/2020 | nd | nd | 12/05/2020 | 2,30E+03 | 2,74E+03 |
| 20/05/2020 | 2,31E+03 | nd | 26/05/2020 | nd | nd | 19/05/2020 | 2,34E+03 | 2,70E+03 |
| 26/05/2020 | nd | nd | 02/06/2020 | nd | nd | 26/05/2020 | nd | nd |
| 02/06/2020 | nd | nd | 09/06/2020 | nd | 2,41,E+03 | 02/06/2020 | nd | nd |
| 10/06/2020 | nd | nd | 16/06/2020 | nd | nd | 09/06/2020 | nd | nd |
|  |  |  |  |  |  | 16/06/2020 | nd | nd |

The first evidence of the presence of SARS-CoV-2 in wastewater samples from Milan was reported in December 2019 (La Rosa et al., 2021), and wastewater was also positive in February 2020, on investigating the gene ORF1ab (La Rosa et al., 2020). In our study, two WWTPs in Milan were investigated; in the first (WWTP1), sewage was monitored from the beginning of March until mid-June 2020, with a peak of SARS-CoV-2 (1.47 E+04 copies/L) on 26 March, followed by a decline (Table 4). The second WWTP (WWTP 2) showed a progressive decline of SARS-CoV-2 concentrations until mid-May when it became undetectable (Table 4). In mid-June, SARS-CoV-2 was not detectable in any of the wastewater samples in Lombardy.

**Table 4.** SARS-CoV-2 in wastewater in the municipality of Milan. Viral targets were two different portions of the nucleocapsid (N) gene, N1 and N3 (copies/L).

| Milano WWTP 1 |  |  | Milano WWTP 2 |  |  |
| --- | --- | --- | --- | --- | --- |
| *Sampling dates* | *N1 copies/L* | *N3 copies/L* | *Sampling dates* | *N1 copies/L* | *N3 copies/L* |
| 03/03/2020 | 5,23E+03 | 2,58E+03 |  |  |  |
| 26/03/2020 | 1,47E+04 | 3,24E+04 |  |  |  |
| 31/03/2020 | 1,39E+04 | 1,43E+04 | 31/03/2020 | 9,19E+03 | 2,31E+03 |
| 07/04/2020 | 7,34E+03 | 4,93E+03 | 07/04/2020 | 2,10E+03 | 2,04E+03 |
| 14/04/2020 | 1,92E+03 | 6,29E+03 | 14/04/2020 | 4,72E+03 | 3,10E+03 |
| 21/04/2020 | 1,73E+03 | 2,55E+03 | 21/04/2020 | nd | 1,42E+03 |
| 28/04/2020 | 2,81E+03 | 2,44E+03 | 28/04/2020 | 1,30E+03 | nd |
| 06/05/2020 | 3,25E+03 | nd | 05/05/2020 | nd | nd |
| 12/05/2020 | nd | 1,68E+03 | 12/05/2020 | nd | nd |
| 19/05/2020 | nd | nd | 19/05/2020 | 1,32E+03 | 2,89E+03 |
| 26/05/2020 | 4,69E+03 | 2,32E+03 | 26/05/2020 | nd | nd |
| 31/05/2020 | nd | nd | 02/06/2020 | nd | nd |
| 09/06/2020 | nd | nd | 09/06/2020 | nd | nd |
| 14/06/2020 | nd | nd | 16/06/2020 | nd | nd |

To the best of our knowledge, the present study reports the largest number of positive wastewater samples (61%) ever found in wastewater monitoring studies. This well reflects the incidence of SARS-CoV-2 infections in the area investigated, which was probably the most affected worldwide in March-April 2020. Samples from the Netherlands and in Massachusetts in March 2020, tested using the same N gene targets, gave SARS-CoV-2 RNA levels up to 2.2 E+03 copies/mL (Medema et al., 2020), and to 3 E+02 copies/mL (F. Wu et al., 2020), respectively. Randazzo et al., 2020 reported results from six cities in the region of Murcia, Spain, with a lower circulation of SARS-CoV-2 and a lower incidence of COVID-19. Ten cities were also investigated in Germany in April 2020 and had lower concentrations of SARS-CoV-2 (2-20 copies/mL) than Italy, but they used a PCR assay targeting two different genes, M and RdRP (Westhaus et al., 2021). In the Czech Republic, 33 WWTPs were investigated in April-June 2020, but only 27% of WWTPs tested positive to SARS-CoV-2 (Mlejnkova et al., 2020).

Other studies investigated temporal trends of SARS-CoV-2 in wastewater, in February-March and May-June, in Brisbane (Australia) (Ahmed et al., 2021), and in Southern Nevada (USA) (Gerrity et al., 2021) testing N1, N2 and E genes. Further monitoring has been done in Japan (Haramoto et al., 2020; Hata et al., 2021), in the United Arab Emirates (Albastaki et al., 2021; Hasan et al., 2021) and in the USA, in Louisiana (Sherchan et al., 2020) and Southeastern Virginia (Gonzalez et al., 2020). Generally in these studies, fewer wastewater samples tested positive for SARS-CoV-2 and concentrations were lower than those observed in Lombardy in the present study.

### 3.4 Quantification of SARS-CoV-2 RNA in wastewater samples

To compare the loads of SARS-CoV-2 among the populations in this study, the viral copies/L detected in wastewater samples were multiplied by the flow rate of each WWTP (m^3^ /day) and the obtained loads (copies/day) were normalized to the number of inhabitants served by each WWTP (Table 5). The N-gene, of which we amplified two portions (N1 and N3 assays), yields similar quantitative results, although there were more positive samples with the N1 fragment, as in previous studies (Medema et al., 2020; Randazzo et al., 2020; F. Wu et al., 2020).

**Table 5.** Viral loads (RNA copies/day/1,000 people) of SARS-CoV-2 in the cities investigated in March-June 2020.

| *Viral load (RNA copies/day/1000 people)* |  |  |  |  |  |  |  |  |  |
| --- | --- | --- | --- | --- | --- | --- | --- | --- | --- |
| *Dates* | *Bergamo* | *Brembate* | *Ranica* | *Brescia* | *Cremona* | *Crema* | *Lodi* | *Milan 1* | *Milan 2* |
| March 30-31 | 3,40E+09 | 9,28E+10 | 1,25E+10 | 1,24E+10 | 1,95E+09 | 2,23E+09 | 1,97E+10 | 2,26E+09 | 3,21E+09 |
| April 1-15 | 2,10E+09 | 2,43E+10 | 8,79E+09 | 3,08E+09 | 2,60E+09 | 4,03E+09 | 2,74E+09 | 8,23E+08 | 1,21E+09 |
| April 16-30 | 7,46E+08 | 4,25E+09 | 3,60E+09 | 6,40E+08 | 6,35E+08 | 6,90E+08 | 3,95E+09 | 6,01E+08 | 7,25E+08 |
| May 1-15 | 4,42E+08 | 2,01E+09 | 3,64E+09 | 0,00E+00 | 0,00E+00 | 4,88E+08 | 4,44E+08 | 4,89E+08 | 0,00E+00 |
| May 16-30 | 0,00E+00 | 1,37E+09 | 2,17E+09 | 3,03E+08 | 4,96E+08 | 0,00E+00 | 3,79E+08 | 3,56E+08 | 3,07E+08 |
| June 1-15 | 0,00E+00 | 3,80E+08 | 1,99E+09 | 5,66E+08 | 0,00E+00 | 6,90E+08 | 0,00E+00 | 0,00E+00 | 0,00E+00 |

In the period end of March - early April 2020, the overall viral load of SARS-CoV-2 that reflected the largest numbers of symptomatic, asymptomatic and paucisymptomatic individuals shedding the virus ranged from 9.3 E+10 copies/day/1,000 people to 8.2E+8 copies/day/1,000 people (Table 5). The load of SARS-CoV-2 RNA copies/day/1,000 people was highest in Brembate (9.28+10), Lodi (1.97E+10), Ranica (1.25E+10), Brescia (1.24E+10) and Bergamo (3.4E+09) in the first week of the study. This is in agreement with the COVID-19 hotspots in March 2020 that were seen exactly in these areas (Bergamo, Brescia and Lodi provinces) as described above. In May, the viral load decreased in all the areas, though low levels persisted until the beginning of June in some cities (Brembate, Ranica, Brescia and Crema). Viral loads were also used in a recent study in Germany, in the Frankfurt metropolitan area, to assess temporal trends of SARS-CoV-2 from April to August 2020 (Agrawal et al., 2021).

### 3.5 Correlation of SARS-CoV-2 RNA detection in wastewater samples and in respiratory samples (active cases)

SARS-CoV-2 RNA levels in wastewater were compared to other epidemiological indicators, such as the number of COVID-19 infections recorded in the corresponding area (Regione Lombardia, 2020). Clearly, virus level in wastewater should not be compared with the number of “new” cases, but with the “total” number of active cases in the population. Total active infections were calculated in all the municipalities served by each WWTP by considering as “active” those diagnosed in the past 4 weeks before the date of sampling, as described above.

Figure 2 and 3 report the correlation among RNA viral loads of SARS-CoV-2 in wastewater and COVID-19 total active cases in the population in the same areas. In general, similar trends were observed, with a pattern of decrease of SARS-CoV-2 in wastewater samples comparable to the decline of the active cases in the population. This very likely reflected the effect of the lock-down at the beginning of March in Lombardy. However, in some cities the trend of the curve of the virus in wastewater seems to anticipate the curve of the number of active cases, pointing to a possible predictive capacity of this indicator for the development of the pandemic, as suggested in other studies (Agrawal et al., 2021; Saguti et al., 2021; Trottier et al., 2020). For instance, in Ranica (Figure 2) an increase of SARS-CoV-2 in wastewater samples was observed in mid-April and early May, followed by an increase of active cases two weeks later.

**Figure 2.**
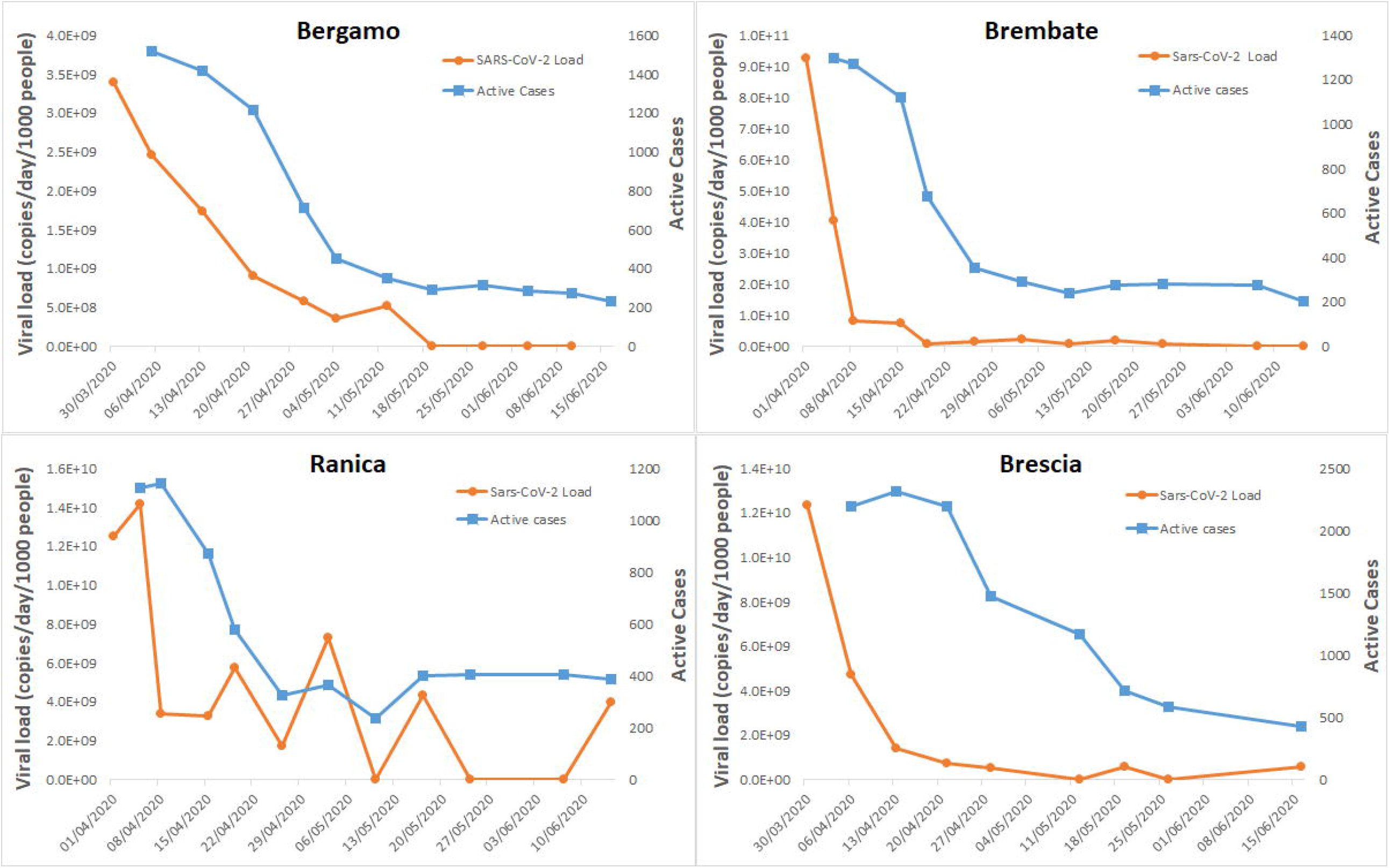
Correlations of SARS-CoV-2 RNA viral loads in wastewater samples and active cases in northwestern Lombardy (Bergamo and Brescia provinces).

**Figure 3.**
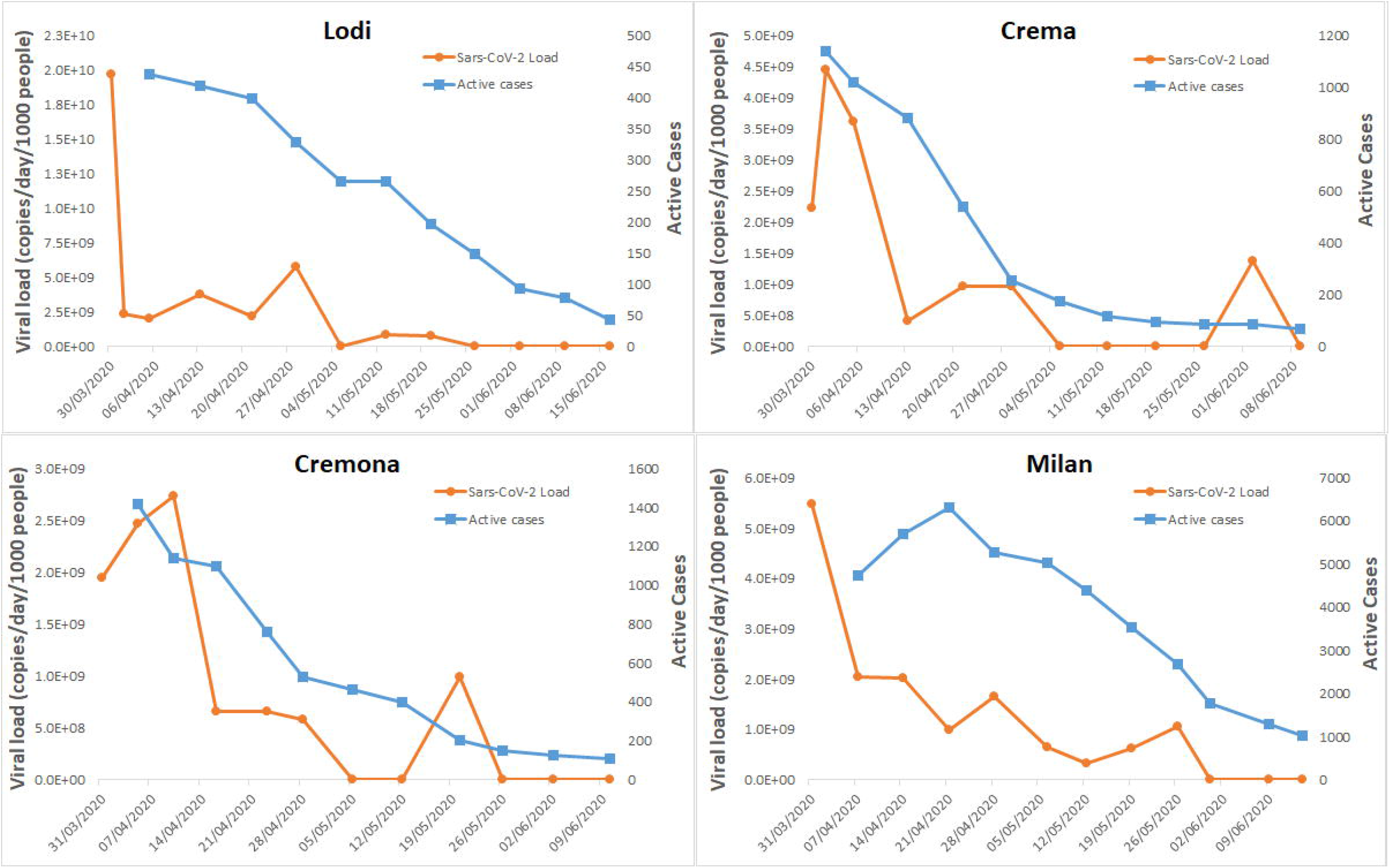
Correlations of SARS-CoV-2 RNA viral loads in wastewater samples and active cases in south Lombardy (Lodi, Crema and Cremona) and in Milan.

### 3.6 Gaps and future perspectives

Several studies, including the present, have demonstrated that monitoring SARS-CoV-2 RNA in wastewater can depict the epidemic curve of COVID-19. Analytical methods set up have been performed promptly, and several useful applications are now available in the literature. Nevertheless, further research is still needed to close some knowledge gaps, related especially to the comparability of the protocols for monitoring SARS-CoV-2 in wastewater, the quality controls and data interpretation.

Several different analytical protocols have been adopted so far (Ahmed et al., 2020c), and they need to be optimized and standardized (Kitajima et al., 2020; Michael-Kordatou et al., 2020) using common quality control procedures. Different external controls, including human coronavirus surrogates, have been tested so far, giving a wide range of recoveries (Alygizakis et al., 2021). The European Commission recently released a first recommendation for a common approach to establish systematic surveillance of SARS-CoV-2 in wastewater (EU, 2021 a), which is the result of a common joint action to share information (European Commission, 2021 b).

In terms of interpreting data, we proposed here the calculation of viral loads and normalization to the population investigated to facilitate comparison between studies. Results from wastewater should also be related to the existing epidemiological indicators. This was an explorative study, and wastewater data were correlated only with the total number of active cases. However, it would be interesting to work out correlations with other epidemiological indicators, such as hospitalization, intensive care demand and death, using also finer statistical tools. This could help make better use of the results from wastewater and valorize the ability of WBE to track infections from symptomatic and asymptomatic individuals in an entire community.

Another possibility lies in estimating prevalence of the infection from the total number of SARS-CoV-2 RNA copies in wastewater by considering the number of viral copies shed in stool by infected individuals (Ahmed et al., 2020a). This estimation was attempted by F. Wu et al., 2020, and indicated a much larger number of infections than the clinical cases recorded. Nevertheless, the uncertainty related to this procedure may be very high due to the wide variability of the load of viral material shed in faeces by asymptomatic, pre-symptomatic and symptomatic cases, the duration of excretion, and limited knowledge of the fate of viral particles in sewer systems. To the best of our knowledge, only one study has investigated SARS-CoV-2 decay in wastewater under different conditions that may simulate the sewer network (Ahmed et al., 2020b). Further information is therefore required to reduce the biases currently associated to relate viral loads to the numbers of cases.

Wastewater monitoring for SARS-CoV-2 may cast light on the real number of cases in a population which – especially during the first phase of the epidemic – was underestimated. In fact, population monitoring through swabbing mainly involved symptomatic individuals, and therefore covered only part of the total individuals infected. Some preliminary data on SARS-CoV-2 in wastewater seems to confirm the initial underestimation of COVID-19 cases in the population. Future research will have to enlarge the case studies and quantitatively assess this phenomenon in Lombardy.

A further application of this surveillance tool that call for more investigation is the identification of virus variants in wastewater and the estimation of their incidence in the population with a single measurement that gives information for an entire community.

## 4. Conclusions

Monitoring SARS-CoV-2 in wastewater may serve as a new surveillance tool to describe the trend of the COVID-19 epidemic in the population. It should be seen as complementary to other epidemiological indicators, but offers the unique capacity to reflect, with a single measurement, the state of the pandemic in a population, and predict the spread of the infection. This indicator can integrate data from the ongoing virological surveillance of COVID-19, providing reliable estimates of the SARS-CoV-2 spread including both symptomatic and asymptomatic individuals, burdening the population-level prevalence of COVID-19 disease, and the real dimension and trends of the epidemic in a population. Therefore, it can be helpful to monitor the effect of public health intervention, such as lock-down on the development of the pandemic.

The surveillance of wastewater samples will also serve as a valuable early warning of the circulation and re-emergence of SARS-CoV-2 infections in the coming months, including the identification of variants. This study will help in establishing a wide surveillance network to follow spatial and temporal profiles of the outbreak.

## Data Availability

the manuscript contains in the text and in the Supplementray material all the data produced.

## Acknowledgements

The authors are grateful to WWTP personnel for sample collection and technical support with information for data analysis. In particular we acknowledge the valid assistance of Uniacque S.p.A, Società SAL srl, A2A Ciclo Idrico S.p.A., Metropolitana Milanese S.p.A., Padania Acque S.p.A.

## Notes

### Competing Interest Statement

The authors have declared no competing interest.

### Clinical Trial

none

### Funding Statement

The study did not had specific funds, it was run with internal resources from our Institutions.

### Author Declarations

The study did not required ethical approval by a committee

