## Supplemental Material for "SARS-CoV-2 RNA in urban wastewater samples to monitor the COVID-19 epidemic in Lombardy, Italy (March – June 2020)"

**Optimization of the concentration procedure**

The following procedures were tested for the concentration of SARS-CoV-2 from wastewater samples:

i) **PEG/dextran procedure (Concentration of 0.5 litre specimen)**

1. Centrifuge the entire sample, in several portions if necessary, for 30 min at 3000 rpm at 4°C. Pool supernatants in a 1 litre flask. Keep the pellets at 4°C.

2. Adjust the pH of the supernatant to neutral (pH 7 –7.5). Measure the volume of the supernatant.

3. To 500 ml of the supernatant, add 39.5 ml of 22% dextran, 287 ml 29% PEG 6000, and 35 ml 5N NaCl. Mix thoroughly and keep in constant agitation for 1 hour at 4°C using a horizontal shaker or magnetic stirrer.

4. Prepare a sterile conical 1 litre separation funnel per sample being evaluated and attach the funnel to a stand. Spread grease on the gliding glass surfaces of the valves but do not obstruct the holes. (Teflon valves do not require smearing). Check water tightness with a small volume of sterile water. Pour the mixture from #3 into the funnels and leave overnight at 4°C.

5. Open the valve with caution. Collect the entire lower layer and the interphase slowly drop-wise, into a sterile tube (usually 5–10 ml per 0.5 litre of original sample).

6. Re-suspend the pellet from #1 into the harvest of #5. Extract with 20% volume of chloroform by shaking vigorously for 1 min. Centrifuge as with faecal suspensions. Collect the upper water phase in a sterile tube

7. Freeze the aliquots of the extracted concentrate at -20°C (-80°C if available) for l future use.

ii) **PEG 8000/NaCl centrifugation procedure (centrifuge at 12000 x g)**
1. Cool the centrifuge to 4 °C.
2. Use pipette (e.g. 25 ml sterile plastic with pipetboy) and transfer 45 ml of wastewater influent in a 50 ml falcon (F1). Balance out all the samples for centrifugation. Prepare two tubes per sample.
3. Centrifuge the samples for 30 min at 4500 x g without brake to remove particulate biomass.
4. Weight 4 g PEG 8000 and 0.9 g sodium chloride into a fresh 50 ml Falcon tube (F2).
5. Transfer 40 ml of supernatant with sterile pipette from F1 to F2.
6. Dissolve PEG/NaCl using a head-over-head shaker up to 15 min at room temperature.
7. Balance the samples for centrifugation using RNase-free water.
8. Before putting F2 into the centrifuge, mark the outer/upper side of the Falcon in order to indicate position of the pellet.
9. Centrifuge the samples for 120 min at 12,000 x g and 4 °C without brake.
10. After centrifugation, decant the sample via the opposing side of the pellet (= marked side up).
11. Centrifuge the Falcons at 12,000 x g for 5 minutes.
12. Aspirate and discard the remaining liquid with a 1,000 µl pipette without touching the pellet.
13. Add 750 µl of trizol to each tube and vortex them for 15 sec to dissolve the pellet.
14. Centrifuge each tube again at 1,000 – 2,000 x g for a few seconds (with brake) to collect all the lysis buffer droplets at the bottom of the falcon.
15. Collect the trizol solutions into a 2 mL tube and store it at -20 until RNA extraction.
iii) **PEG 6000 and 8000/NaCl centrifugation procedure (centrifuge at 3893 x g)**
1. Cool the centrifuge to 4 °C.
2. Use pipette (e.g. 25 ml sterile plastic with pipetboy) and transfer 45 ml of wastewater influent in a 50 ml falcon (F1). Balance out all the samples for centrifugation. Prepare two tubes per sample.
3. Centrifuge the samples for 30 min at 3893x g without brake to remove particulate biomass.
4. Weight 4 g PEG 6000/8000 and 0.9 g sodium chloride into a fresh 50 ml Falcon tube (F2).
5. Transfer 40 ml of supernatant with sterile pipette from F1 to F2.
6. Dissolve PEG/NaCl using a head-over-head shaker up to 15 min at room temperature.
7. Balance the samples for centrifugation using RNase-free water.
8. Centrifuge the samples for 120 min at 3893x g and 4 °C without brake.
9. After centrifugation, decant the sample via the opposing side of the pellet (= marked side up).
10. Centrifuge the Falcons at 3893x g for 5 minutes.
11. Aspirate and discard the remaining liquid with a 1,000 µl pipette without touching the pellet.
12. Add 750 µl of trizol to each tube and vortex them for 15 sec to dissolve the pellet.
13. Centrifuge each tube again at 1,000 – 2,000x g for a few seconds (with brake) to collect all the lysis buffer droplets at the bottom of the falcon.
14. Collect the trizol solutions into a 2 mL tube and store it at -20 until RNA extraction.

**Table S1.** Ct results of N1 and N3 for the different concentration procedure tested. The procedure adopted for analyses is highlighted.

*undet= undetectable, below LOD=1 copy/uL.

| **Concentration method** | **Treated Volume** | **Ct Value**  **N1**  **(duplicate analysis)** | **Mean**  **N1 Ct** | **Ct Value**  **N3 (duplicate analysis)** | **Mean**  **N3 Ct** |
| --- | --- | --- | --- | --- | --- |
| PEG/dextran | 250 mL | undet* | undet* | undet* | undet* |
|  |  | undet* |  | undet* |  |
| PEG 6000/ NaCl centrifugation (3893g) | 80 mL | 36.76 | 36.15 | undet* | undet* |
|  |  | 35.53 |  | undet* |  |
| PEG 8000/ NaCl centrifugation (3893g) | 80 mL | 35.22 | 35.40 | 36.23 | 35.36 |
|  |  | 35.58 |  | 34.5 |  |
| **PEG 8000/ NaCl centrifugation (12000g)** | **80 mL** | **32.31** | **32.54** | **33.63** | **33.59** |
|  |  | **32.77** |  | **33.55** |  |
| PEG 8000/ NaCl centrifugation (12000g) + particulate | 80 mL | 32.1 | 31.30 | 32.36 | 32.40 |
|  |  | 30.59 |  | 32.37 |  |

**Table S2.** Optimization of the volume of extraction on QIAamp MinElute columns. Results from both RT-qPCR and droplet digital PCR (ddPCR).

| **Volume of extraction** | **RT- qPCR (duplicate analysis)** | | **ddPCR (triplicate analysis)** | | |
| --- | --- | --- | --- | --- | --- |
|  | **Mean N1 Ct**  **(SD)** | **Mean N3 Ct**  **(SD)** | **Mean N1 copies/mL**  **(SD)** | | **Mean N2 copies/mL**  **(SD)** |
| Extraction 200 µL | 34.22 (0.23) | 34.04 (1.3) | 393.0 (170) | 245.0 (94) | |
| Extraction 400 µL | 33.80 (0.37) | 34.29 (0.17) | 827.3 (354) | 471.7 (24) | |

**Table S3.** Optimization of the volume of elution from the QIAamp MinElute columns. Results from ddPCR analysis. Results are the means of triplicate analysis with standard deviation (SD).

| **Volume of elution** | **1 step - 100 µL** | | **2 steps - 50 +50 µL** | | **1 step - 60 µL** | |
| --- | --- | --- | --- | --- | --- | --- |
| **Target genes** | **N1** | **N2** | **N1** | **N2** | **N1** | **N2** |
| **Copies/mL** | 134.9 (4.6) | 202.8 (62.6) | 538.5 (254.5) | 236.3 (29.9) | 574.2 (128.3) | 406.8 (151.8) |
